# The LIFE Study: a cross-sectional study protocol for LIfestyle risk Factors for chronic disease across the stagEs of reproductive ageing

**DOI:** 10.64898/2026.01.28.26344893

**Authors:** Laura E Pernoud, Jamie L Noll, Paul A Gardiner, Melinda M Dean, Kathryn M Broadhouse, Meegan A Walker, Hattie H Wright, Anthony Villani, Joseph J Scott, Alexandra P Metse, Mia A Schaumberg

## Abstract

The dynamic physiological and hormonal changes through the menopause transition predispose women to an increased risk of multiple chronic diseases including cardiovascular disease, metabolic disease, depression and dementia. The underlying mechanisms remain unclear, yet it is thought that chronic systemic inflammation and changes to lifestyle behaviors across menopause play important roles. The LIfestyle risk Factors for chronic disease across the stagEs of reproductive ageing (LIFE study) is a cross-sectional study aimed to develop an understanding of how hormonal and lifestyle factor differences across pre, peri and postmenopause influence chronic systemic inflammation visceral adiposity, cognitive function and sleep health. A total of 165 women aged between 40-65 years have been recruited and classified into pre, peri or postmenopausal groups. Body composition measures and blood glucose samples were collected. Sleep and physical activity were objectively measured using activPAL4 and ActiGraph GT9X link accelerometer over 7 days. Participants were also provided with a sleep log diary. Physical function was assessed using the Short Physical Performance Battery. Cognitive function was evaluated using Addenbrooke’s Cognitive Examination-III and Cambridge Neuropsychological Test Automated Battery. Participants completed a series of questionnaires including the Depression, Anxiety, and Stress Scale-21, RuSATED, Berlin Questionnaire, Insomnia Severity Index, Activities-specific Balance Confidence Scale, and the Australian Eating Survey. The LIFE study aims to firstly determine how differences in lifestyle behaviors (physical activity, diet and sleep) across menopause influence chronic systemic inflammation and visceral adiposity. Secondly, to determine the association between chronic systemic inflammation, visceral adiposity, and cognitive function, elucidating the relationship between lifestyle behaviors and menopausal symptoms. Collectively this will provide an understanding and characterization of lifestyle behaviors and links to inflammatory markers, cognition, mental health and sleep health in pre, peri and postmenopausal women that will inform targeted strategies to improve long-term, wellbeing, heart, brain, and metabolic health.

## INTRODUCTION

The menopause transition is defined as a dynamic period of physiological change, ending in complete ovarian failure and cessation of reproductive function (1). The rapid decline in the production and secretion of ovarian hormones, particularly, oestrogen, is associated with changes in mental health, cognitive, cardiovascular, immune and metabolic functions, and unfavourable changes in body composition (2–5). These changes predispose postmenopausal women to an increased risk of multiple diseases including osteoporosis, hypertension, cardiovascular disease, metabolic disease, depression and dementia (6–9).

Lifestyle-related factors, particularly at midlife, play a critical role in the reduction of chronic disease risk and inflammation. Factors such as weight gain, increased sedentary behavior, poorer sleep quality and diet all contribute to an increase in low-grade inflammation (10, 11). It is this chronic systemic inflammation that plays an important role in the etiology of many of the diseases known to increase in prevalence in women after menopause (2, 6, 12, 13). Currently, the underlying mechanisms to explain this increase in prevalence in postmenopause and the link to systemic inflammation is poorly understood and little is known about the complex biological changes that occur during the menopausal transition that may contribute to the development of chronic disease.

Incidence of overweight and obesity is increased in postmenopausal women compared to premenopausal women via multifactorial influences including reduced oestrogen production, reduced energy expenditure and lower basal metabolic rate (14, 15). The withdrawal of oestrogen, specifically 17β-oestradiol, drives a shift in the deposition of adipose tissue from subcutaneous gluteal femoral regions to ectopic sites around metabolically active visceral organs, including the liver, pancreas, and skeletal muscle (3, 13, 15, 16). As a result, circulating markers of inflammation, glucose, and lipids become elevated and instigate disruptions in insulin production, secretion and action, mechanisms which underlie the development of chronic disease (4, 17, 18). These changes to body composition may explain the increase in chronic inflammation in postmenopausal women, and subsequent increase in disease prevalence (19, 20).

Adipose tissue secretes hormones and inflammatory cytokines which play an integral role alongside oestrogen in the regulation of energy intake, expenditure and storage (14, 21). In overweight and obesity, adipose tissue dysfunction occurs, where an overproduction of hormones and inflammatory cytokines such as leptin, interleukin-1β (IL-1β), interleukin-6 (IL-6), tumour necrosis factor-a (TNF-α), among others, contributes to a state of low-grade inflammation (22–25).

For women approaching menopause, the confounding effect of excess body fat, alongside declining ovarian function and withdrawal of oestrogen, perpetuates chronic inflammation and further heightens the risk of chronic disease (26, 27). Studies have shown a strong link between increased proinflammatory cytokine activity and risk of cardiovascular disease, metabolic syndrome and type 2 diabetes (20, 28). Chronic elevations in proinflammatory cytokines are also associated with alterations in cognitive function (29, 30). The depletion of ovarian hormones during postmenopause has been considered a risk factor for accelerated cognitive decline in women (31, 32). However, no studies have concurrently investigated the underlying mechanisms between mental health status, cognition, inflammation and increased adiposity that ensue post-menopause.

Very few studies in the literature have compared markers of systemic inflammation across menopause, with significant limitations in the measurement of body composition, classification of menopausal stage, and lack of hormone verification of menopause stage (2, 9, 13, 33, 34). Furthermore, research investigating the relationship between lifestyle factors (e.g., diet, physical activity, sleep) and pro-inflammatory markers and the impact on chronic disease progression is sparse. By characterising lifestyle behaviors across menopause, the LIfestyle risk factors for chronic disease across the stagEs of reproductive ageing (LIFE) study will inform targeted strategies to improve long-term heart, brain, and metabolic health. Understanding the relationship between menopause, inflammation, and body composition, will direct future lifestyle interventions for older women at risk of obesity-related chronic disease. Finally, investigating the underlying mechanisms linking unfavourable changes in body composition, chronic inflammation, mental health and cognitive function will further our understanding of inflammatory responses to menopause, and how these changes relate to wellbeing, cognitive decline and increased risk of dementia.

## STUDY AIMS AND HYPOTHESES

The primary aim of the LIFE study is to firstly, characterize lifestyle behaviors such as sleep, physical activity, dietary intake and pro-inflammatory markers such as leptin, IL-1β, IL-6 and TNF-α, between BMI-matched, pre, peri and postmenopausal women. The secondary aim is to identify whether the differences/changes in lifestyle factors and pro-inflammatory cytokines in pre, peri and postmenopausal women are associated with visceral adiposity. The third aim is to investigate whether differences/changes in pro-inflammatory cytokines and visceral adiposity between pre, peri and postmenopausal women are associated with wellbeing and cognitive function. The final aim is to investigate the association between lifestyle behaviors (sleep, diet, sedentary behavior, physical activity) and menopausal symptoms.

It is hypothesized that firstly, there will be distinct lifestyle behaviors and 24-hour time-use characteristics between menopausal groups. Secondly, that there will be significant and distinct relationships between inflammation markers and lifestyle measures between each group. Thirdly, that increases in visceral adiposity will correlate with elevated chronic systemic inflammation, and lower wellbeing scores and cognitive performance in peri and postmenopausal women when compared to premenopausal women. Finally, it is hypothesized that there will be a negative association between lifestyle behaviors and menopausal symptoms across all groups.

## STUDY DESIGN

The LIFE study is a cross-sectional study conducted at the host institution. Recruitment commenced in October 2022 and data collection was completed in May 2024. Data analysis is ongoing and is expected to be completed by 2026. Ethical approval was received from the relevant University Human Research Ethics Committee (ethics approval number #S221718) prior to the commencement of the research project. Participants were recruited, screened for menopausal status in the first assessment session and then underwent two subsequent assessment sessions to collect demographic and lifestyle data.

### Sample size calculation

A sample size estimate was calculated using G*Power (version 3.1.9.4). As the primary outcomes of the LIFE study are to characterise lifestyle behaviors and investigate associations between inflammation markers across pre, peri and postmenopausal groups an effect size of 0.30 was pooled from previous published studies investigating differences in IL-6 concentrations between pre and postmenopausal women (3, 35, 36). Using an alpha level of 0.05, and a power level of 95%, it was determined that a total sample size of n=175 was required for ANCOVA analysis of 3 groups and 3 covariates.

### Participants and study setting

Participants were eligible if they identified as a cisgendered woman, or non-binary (female sex at birth and gender identity is non-binary), were aged between 40-65 years, had a body mass index (BMI) 18.5-39.9 kg/m^2^, were able and willing to provide informed consent, and lived locally to, or willing to travel to the study location. Participants were excluded if they were pregnant, or breastfeeding, were using exogenous hormones such as hormonal replacement therapy, or oral contraceptives, received treatment or surgically induced menopause (e.g. oophorectomy, hysterectomy), and had a diagnosis of a hormonal or ovarian disorder such as polycystic ovarian syndrome, or hypothalamic amenorrhea. To ensure an equal distribution of participants across the three groups (pre, peri and postmenopausal), once the target number of 58 participants was achieved in the postmenopausal group, recruitment for postmenopausal women ceased. Complete details of eligibility and exclusion criteria are provided in Supplementary table 1.

### Recruitment sources

Recruitment was undertaken using several recruitment methods, including but not limited to, disseminating study material across the study location campus, women’s health clinics, and hospitals within the local community, online social media platforms, television and radio feature segments, community announcement boards, local community groups and participant word-of-mouth.

### Screening and consent

Interested participants submitted their details, including name, contact information, residential postcode, date of birth and self-classified menopause status, via an online form located on the LIFE study-specific webpage through the host institution. Upon receiving an expression of interest, researchers provided participants with an information consent form via email, along with a link to an online screening form to assess eligibility. Eligible participants were contacted to discuss study procedures through a telephone-based interview and invited to participate in the study. Once participants had agreed to participate, they were scheduled for two, in-person assessment sessions, conducted one week apart. A confirmation email was sent with details of both assessment sessions, and a map to the testing location. Ineligible participants were notified and referred to alternate suitable research studies when possible. The flow of participants through the study, including reasons for exclusion, are shown in Figure 1.

**Figure 1.**
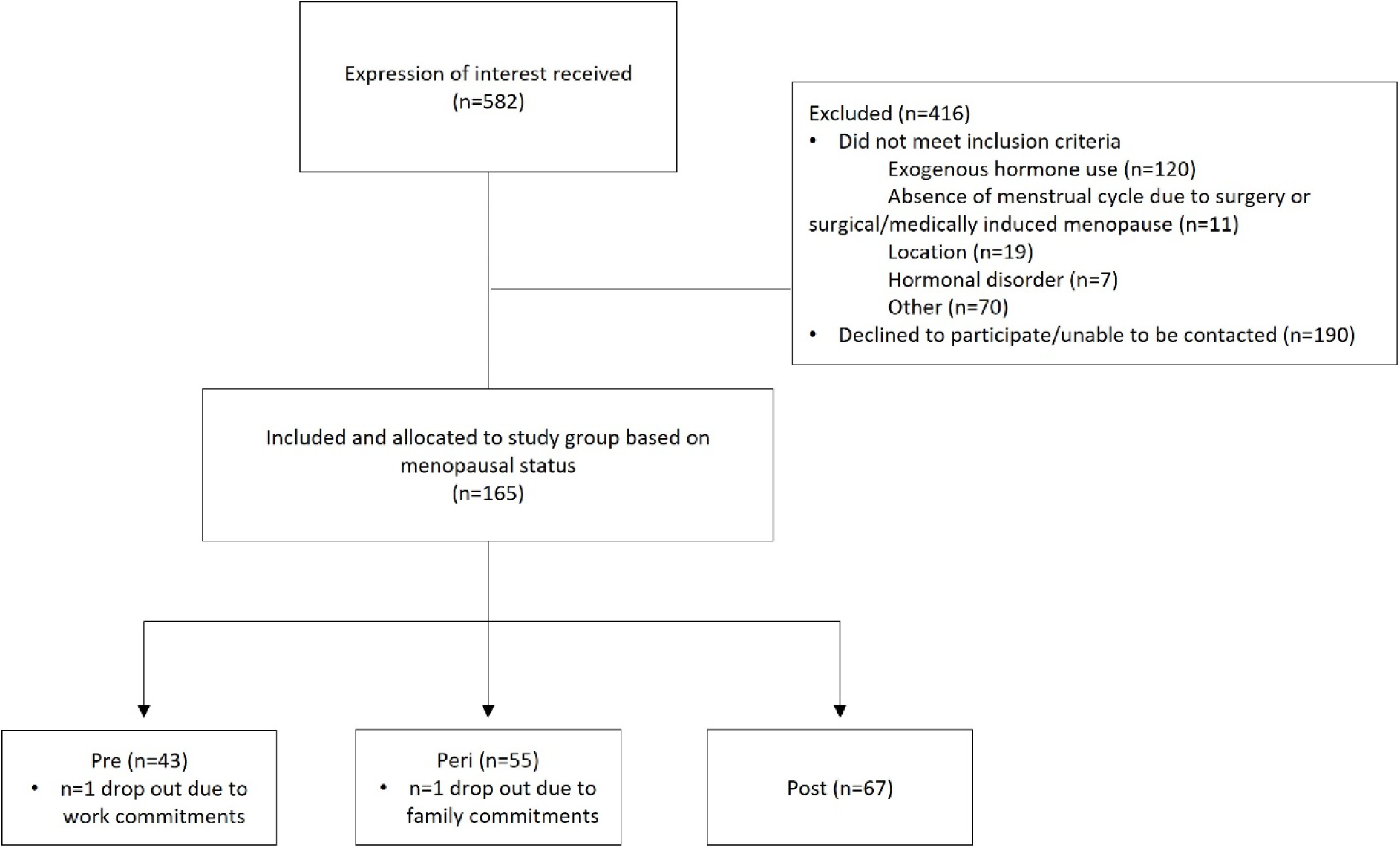
Participant flow diagram. Interested participants submitted their details via an online form located on the LIFE study-specific webpage through the host institution. Upon receiving an expression of interest, researchers provided participants with an information consent form via email, along with a link to an online screening form to assess eligibility and menopausal status. Allocation of eligible participants, number of dropouts and reason are shown above.

## STUDY PROTOCOL

### First data collection: menopausal classification

Two to three days prior to the initial in-person assessment, a reminder email was sent to participants confirming the date, time, and pre-testing preparation. They were asked to complete the Reproductive Ageing in Women questionnaire (RAW) via the online platform, Qualtrics. Based on their responses, participants were classified into pre, peri or postmenopausal groups. Participants with a regular menstrual cycle mostly the same length apart, and no presence of hot flushes/flashes were classified as premenopausal. Participants were classified as perimenopausal if they had either: a regular menstrual cycle accompanied by the presence of hot flushes/flashes, or a variable menstrual cycle (defined as a difference of 7 or more days in cycle length between cycles), and/or the presence of hot flushes. Participants who had not experienced a menstrual cycle for 12 months or more due to physiological menopause were classified as postmenopausal. In addition, participants were asked whether they had noticed any changes in menstrual bleeding including heaviness of flow, and/or changes in menstrual pain (increase or decrease). They were also asked if they were currently taking or using medications, supplements (including vitamins), or any alternative therapies (e.g., acupuncture, or yoga) to manage menopausal related symptoms. The RAW questionnaire took approximately 30 minutes to complete and included 19 items related to menstrual cycle length and characteristics, hormonal contraception use, time since the final menstrual period (for postmenopausal participants), cause of menstrual cycle cessation, and use of menopausal treatments. The Menopause-specific Quality of Life Questionnaire (MENQOL) (37) Audit-C (38), Active Australia Survey (39), and the Pittsburgh Sleep Quality Index (PSQI) (40).

### Second data collection: Demographics, blood sampling and lifestyle measures

Participants arrived at the study location campus between 5:00-9:00am wearing enclosed shoes and light, non-restrictive clothing with no zips, underwires, or metal and having fasted (≥8 hours). After providing written consent, participants completed a brief questionnaire collecting contact details, next of kin and local GP information, medication use, and medical history. Medical history included details regarding prior radiation exposure to assess whether participants were permitted to undergo bone mineral density and body composition scans.

Demographic and lifestyle measures were then collected. Measurements included body mass, standing height, waist and hip circumferences, and assessments of bone mineral density and body composition using dual x-ray absorptiometry (DXA), bioelectrical impedance spectroscopy (BIS) and air displacement plethysmography (ADP). Participants were then fitted with an activPAL4 device (worn on the thigh), and an ActiGraph GT9X Link watch (worn on their non-dominant wrist) to monitor physical activity and sleep objectively over a period of seven-days. Participants were also provided with a sleep diary and instructed how to complete it. Blood glucose was measured via capillary whole blood collected from the medial or lateral aspect of the third, fourth or fifth finger using a sterile lancet device.

Capillary blood was then analysed instantly (Accu Chek Instant S Meter Kit) to determine fasting blood glucose levels. Blood sample collection for pre and perimenopausal women with regular menstrual cycles was scheduled between days 2-7 of their menstrual cycle (41). Perimenopausal women who had not experienced a regular cycle in the 3 months prior, and postmenopausal women were scheduled on a day that was most convenient for the participant.

### Third data collection: Physical and cognitive assessments

The third data collection session took place face-to-face at the study location campus, at least 6 days after the first assessment (average 7.6 ± 2.0 days, range: 6-23 days) in a non-fasted state, with participants refraining from caffeine or alcohol ≥4 hours and strenuous exercise ≥12 hours before arrival. Participants returned the sleep diary and objective monitoring devices, after which seated blood pressure was measured. Physical function was assessed using grip strength, 30-second sit-to-stand, balance tests (semi-tandem, tandem, single-leg), timed-up-and-go, and a 6-minute walk test. Cognitive function was assessed with the Addenbrooke’s cognitive examination (ACE-III) (42), and the Cambridge Neuropsychological Test Automated Battery (CANTAB) (43). Participants also completed a series of validated questionnaires including the Depression, Anxiety, and Stress Scale-21 items (DASS-21) (44), RuSATED (45), Berlin Questionnaire (46), Insomnia Severity Index (ISI) (47), pre-sleep arousal scale (PSAS) (48), Activities-specific Balance Confidence Scale (ABC) (49), and the Australian Eating Survey (AES). The study protocol is detailed in Figure 2.

**Figure 2.**
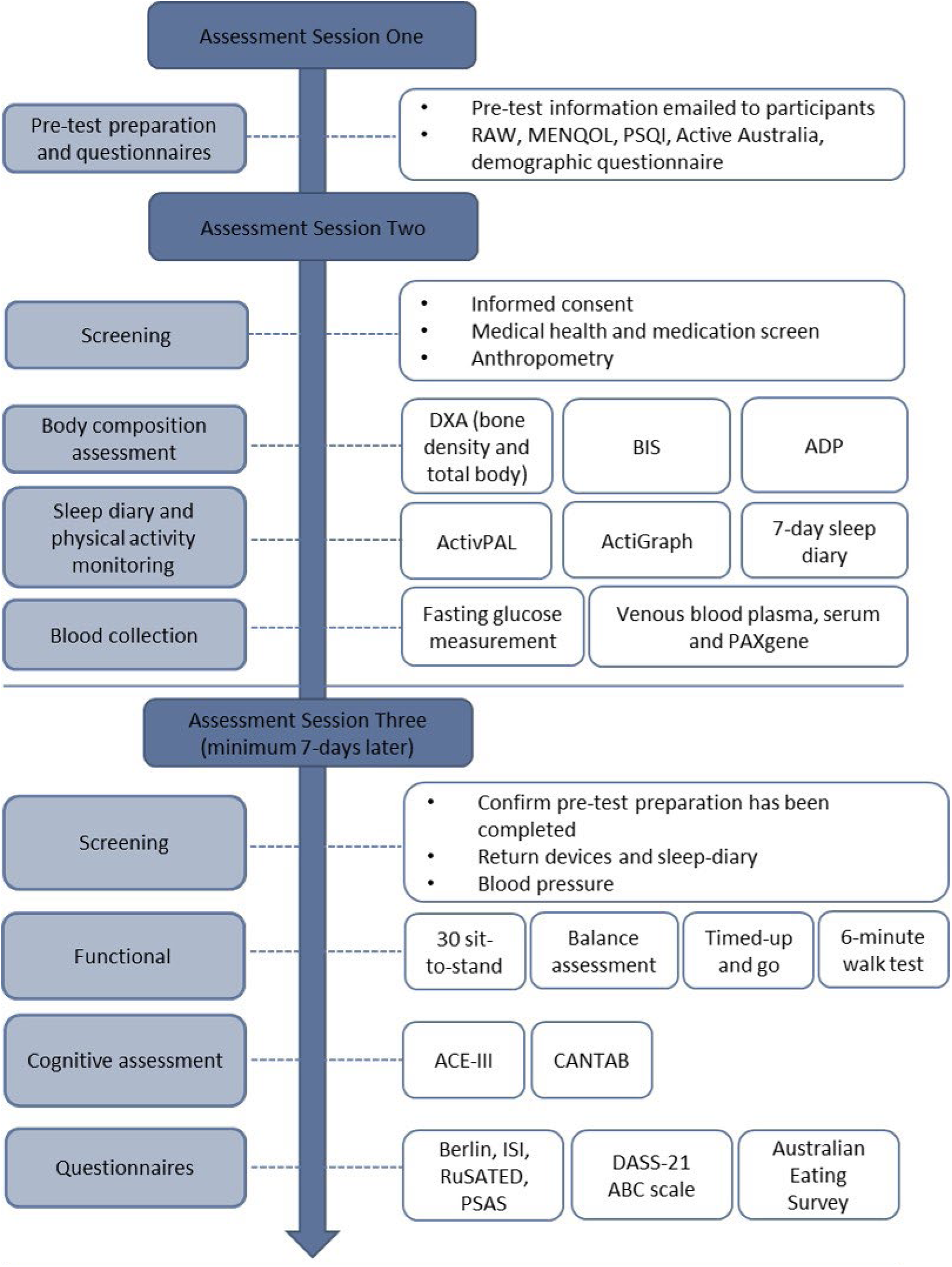
Flow diagram of LIFE study protocol. RAW: Reproductive ageing in women questionnaire, MENQOL: Menopause-specific quality of life questionnaire, PSQI: Pittsburgh sleep quality index, DXA: Dual x-ray absorptiometry, BIS: Bioelectrical impedance spectroscopy, ADP: Air displacement plethysmography, ACE-III: Addenbrooke’s cognitive examination-III, CANTAB: Cambridge Automated Neuropsychological Test Automated Battery, ISI: Insomnia severity index, DASS-21: Depression Anxiety Stress Scale, ABC: Activities-Specific Balance Confidence scale.

## DATA COLLECTION

Data collection procedures used across the three assessment sessions are detailed below. Table 1 provides a summary of the objective and questionnaire-based measures along with their intended purpose in the study (e.g., primary outcomes, secondary outcome).

**Table 1.** Summary of study outcomes and intended purposes.

| Component | Key constructs | Measures |  |
| --- | --- | --- | --- |
|  |  | Primary/secondary outcomes | Other measures |
| **Fasting blood draw** | Inflammatory markers<br>Neurotrophic factors<br>Genetic risk factors<br>Bone markers<br>Reproductive hormones<br>Blood glucose | TNF-α, IL-1β, IL-6, IL-8, IL-10, MCP-1, IFN-γ,<br>Leptin, Adiponectin,<br>VEGF-A, BDNF<br>FSH, E2 | ApoE4<br>OPN, OPG<br>Capillary blood glucose |
| **Anthropometric** | Body weight and body composition<br>4C model of body composition | Weight, WC, WHR, DXA assessed anteroposterior spine BMD, left femur BMD, total FM, LM, total BF %, DXA software estimated VAT<br>BIS assessed TBW, and ADP assessed TBD.<br>The 4C model of body composition combined total BMC, TBW, TBD and body weight to calculate the most accurate measure of total body fat | Height, HC, BMI |
| **Vascular** | Blood pressure and heart rate |  | Seated systolic blood pressure, diastolic blood pressure and heart rate |
| **CANTAB** | Visual learning and new memory<br>Verbal learning and new memory<br>Attention and psychomotor speed | PAL total errors (adjusted), PAL total errors 12 shapes (adjusted)<br>VRM immediate recognition total correct (adjusted), delayed recognition total correct (adjusted), total score immediate recall (adjusted), free recall double-scored, free recall percentage.<br>RTI simple median reaction time, RTI simple median movement time & RTI median 5-choice reaction time, RTI median 5-choice movement time | PAL total errors, PAL total errors 12 patterns<br>Immediate recognition incorrect rejections, immediate recognition total errors, delayed recognition incorrect rejections, delayed recognition total errors.<br>RTI simple error score (all), RTI total error score (5-choice) |
|  | Visual matching ability and visual recognition memory<br>Working memory | DMS percent correct, DMS median correct latency<br>SWM between errors, SWM between errors (12 boxes),<br>SWM strategy (6-12 boxes) | DMS total errors, DMS total correct (12s delay)<br>SWM total errors, SWM total errors (12 boxes) |
| **Cognitive function** (ACE-III) | Global cognition | Attention, memory, fluency, language, visuospatial | MMSE Score |
| **Self-reported Sleep**<br>PSQI<br>Consensus Sleep Diary<br>RU-SATED<br>ISI<br>PSAS<br>BQ<br>Self-ME | Subjective sleep quality<br>Multidimensional sleep health<br>Insomnia symptoms<br>Cognitive and somatic arousal<br>Sleep disordered breathing<br>Chronotype preference | Overall sleep quality (past month)<br>Nightly: sleep onset latency, wakefulness after initial sleep onset, total sleep time, total time spent in bed, sleep efficiency, and sleep satisfaction, Sleep regularity, timing, satisfaction, duration and efficiency and daytime alertness.<br>Symptoms of cognitive and somatic arousal experienced at bedtime | Duration of sleep, Sleep disturbance, sleep latency, day dysfunction due to sleepiness, sleep efficiency<br>Sleep timing and regularity.<br>Nature, severity and impact of insomnia symptoms (past 2 weeks).<br>Risk of Obstructive Sleep Apnea<br>Chronotype preference (morning or evening person) |
| **Self-reported Physical activity** (Active Australia) | Physical activity |  | Number of sessions of physical activity, total time spent in each activity (minutes), average time spent in each activity (minutes) (past week) |
| **Device-based monitoring** (thigh-worn activPAL4) | Physical activity<br>Sedentary behavior | Standing time, stepping time, LPA, MVPA<br>Overall sitting time (m), overall prolonged sitting time, ($\geq 30$ min), activity score (MET.h) | |
| **Device-based monitoring** (wrist-worn ActiGraph) | Sleep<br>Physical activity | TST, SE<br>Sedentary behaviors, LPA, MVPA | SOL, WASO, sleep fragmentation, sleep regularity, sleep timing |
| **Mental Health**<br>DASS-21 | Depression<br>Anxiety<br>Stress |  | Dysphoria, hopelessness, devaluation of life, self-deprecation, lack of interest, anhedonia, and inertia |
|  | General Psychological Distress |  | Autonomic arousal, skeletal muscle effects, situational anxiety, subjective experience of anxious affect<br>Difficulty relaxing, nervous arousal, easily upset/agitated, irritable/overreactive and impatience |
| **Functional performance** | Functional ability |  | Hand grip strength, semi-tandem, tandem and single leg balance assessment, 30-second sit-to-stand, timed-up-and go, 6-minute walk test ABC scale |
| **Habitual diet**<br>AES | Dietary intake |  | Daily energy and nutrient intake (macro / micronutrients) over the preceding 6 months. |
| **Menopause status**<br>RAW | Menopause classification | Menstrual history, changes in menstrual cycle, current menstrual status, vasomotor symptoms, hormone therapy or supplements | Menopausal symptoms (physical, psychosocial, sexual) (past month) |

### Classification of menopause status and symptoms

The RAW questionnaire contained questions to assist in classifying the participants menopause status according to the Stages of Reproductive Aging Workshop (STRAW+10) criteria (50). To classify menopause status, participants were asked questions such as current menstrual cycle status and regularity of their menstrual cycle. Length of time spent with menstrual cycle variability or time since the final menstrual period, and cause of menstrual cycle cessation was also reported. In addition, participants were asked whether they had noticed any changes in menstrual bleeding including heaviness of flow, and/or changes in menstrual pain (increase or decrease). Hormonal contraceptive use, hormonal therapy, and associated symptoms were recorded. This questionnaire was developed specifically for the LIFE study, and its measurement properties are currently being evaluated for reliability and validity.

As part of the RAW questionnaire, participants completed the MENQOL, which is self-administered and consists of a total of 29 items in a Likert-scale format. Participants responded to questions in each of the four domains of menopausal symptoms, as experienced over the last month: vasomotor (items 1–3), psychosocial (items 4–10), physical (items 11–26), and sexual (items 27–29). Participants rated each symptom as present or absent. If present, its level of bothersome was scored on a 0 (not bothersome) to 6 (extremely bothersome) scale. Means were computed for each subscale by dividing the sum of the domain’s items by the number of items within that domain. Non-endorsement of an item is scored a “1” and endorsement a “2,” plus the number of the rating, so that the possible score on any item ranges from one to eight (37).

### Anthropometry measures

<u>Body mass</u> was measured to the nearest 0.1 kg using a set of electronic stand-on scales (AND HW-200GL, A&D Australasia Pty Ltd, Thebarton, South Australia) where participants were asked to wear comfortable light clothing, empty their pockets and remove footwear. Height was measured to the nearest 0.1 cm using a wall mounted stadiometer with the participants head positioned in the Frankfort plane, feet together with their back against the stadiometer. Both measures of weight and height were undertaken in accordance with the International Society for the Advance of Kinathropometry (ISAK) protocols (51).Body mass Index (BMI) was calculated as weight (kg) divided by the square of height (m^2^) (52).

#### Waist and hip circumference

Participants stood in an upright position while waist and hip measurement was conducted. The tape was passed around the; waist: abdomen at the narrowest point between the lower costal and top of the iliac crest; hip: widest point of the gluteals/hips. Measurements were completed in duplicate with the mean value recorded as per previously published protocols (53). *Waist-to-hip ratio* (WHR) was calculated as waist circumference (cm) divided by hip circumference (cm).

### Assessment of body composition and bone mineral density

#### Body composition measures

Dual X-ray absorptiometry (DXA) was used to determine body composition (fat mass, lean body mass, body fat percentage, android and gynoid fat mass and android/gynoid ratio) and bone mineral density (BMD). The DXA system was calibrated with phantoms as per the manufacturers guidelines each day before measurements were taken. Participants were positioned in the centre of the densitometer table with their hips and knees flexed on a supportive block to flatten the lordosis of the lumbar spine. Antero-posterior spine BMD measurements were obtained using the L1-L4 vertebral bodies. For left hip BMD, the measurement was performed after aligning the long axis of the femur with the scanner. A positioning device was used to internally rotate the femur to elongate the femoral neck. Bone mineral density measurements were obtained using the femoral neck, greater trochanter, Ward’s area, intertrochanteric regions, and total hip. Participants were re-positioned to the centre of the densitometer table for the total body scan (54) with their feet placed in a radio-opaque styrofoam block, secured using a Velcro strap around the ankles to maintain a distance of 15cm between the feet (55). Participants positioned their hands laterally against the hips within shaped Styrofoam blocks with a consistent gap of 3cm between the palms and trunk. The arms were held in place with a Velcro strap to minimise any subject movement during the scan (55). This positioning protocol has previously been shown to enhance measurement precision, and minimize typical errors associated with technical and biological variability of DXA measurements of whole and regional body composition when compared against the traditional NHANES protocol (55). All DXA scans were undertaken using the GE Lunar iDXA (Lunar DPX, GE Healthcare, Madison, WI), and were analysed using software (GE encore version 18.0) provided by the manufacturer (GE Healthcare) and according to the manufacturer’s instructions. All scans were conducted by DXA trained and accredited research personnel, at the host institution. Participants wore exercise clothing, and were instructed to remove shoes, hats, and any metal-containing items such as belts, watches, jewellery or coins.

#### Total body water measurements

Bio-electrical impedance spectroscopy (BIS) (Imp™ SFB7) was used to assess total body water (TBW). The BIS was calibrated as per manufacturers guidelines each day before measurements were taken. Participants were supine for a minimum of 10 minutes prior to, and for the duration of the assessment. Two, single tab electrodes (3M Healthcare) were placed on the left hand, next to the wrist joint and on the dorsal surface of the hand proximal to the middle knuckle with at least a 5cm gap between them. Two electrodes were placed on the left ankle, on the dorsal surface of the foot at the level of the protruding bone on the side of the ankle, and the dorsal surface of the foot with at least a 5cm gap between them.

#### Body mass and volume measures

Air Displacement plethysmography (ADP) technique using the BOD POD^®^ (Cosmed, Italy) system was used to measure the subject’s mass and volume via electronic scale and air displacement to derive body density. The BOD POD system was calibrated as per manufacturer’s instructions each day before measurements were taken. Participant mass was measured, and the participant was then required to sit in the chamber for approximately 5 minutes. Thoracic gas volume was predicted. As per operation guidelines, the average of two scans was taken as the result for one assessment. Unless the variance was greater than 150 mL, then a third measure was taken, and the mean used for subsequent analysis. Participants wore exercise clothing and a swimming cap to minimise measurement error.

### Device-based measurement of physical activity, sedentary behavior, and sleep

Device-based objective measures of physical activity, and sleep were collected using the activPAL4 activity monitor (PAL Technologies Limited, Glasglow, UK; 24-hr protocol) worn on the thigh and the ActiGraph GT9X Link (ActiGraph Corporation, Pensacola, FL) worn on the wrist of the non-dominant hand. The devices were worn continuously for seven consecutive days and were removed only for bathing and water-based activities (e.g., swimming).

The activPAL4 was initialized and waterproofed with a plastic sleeve and attached to the participants approximately one-third from the top of the thigh using either standard (3M™ Tegaderm™) or hypo allergenic (Opsite Flexifix Gentle) film. Data was imported and analysed using the PALanalysis v8.11.8.75, CREA Algorithm v1.3. Data was deemed valid if at least one full day of recording was available (≥10 hours wear time). The activPAL4 uses triaxial accelerometry to reliably discriminate periods of upright activity from seated or lying activities (56). The following outcomes will be derived from the ActivPAL4 data: standing time, stepping time, light physical activity (LPA) (cadence ≥75, duration>1m), moderate to vigorous physical activity (MVPA) (cadence≥100, duration>1m) (57), overall sitting time (mins), overall prolonged sitting time (≥30 min), and activity score (MET.h).

The ActiGraph GT9X Link tri-axial accelerometer was initialized with a sampling rate of 90 Hz and was set to begin recording at midnight following the day the participants received the device. ActiGraph data were downloaded using ActiLife software (version 6.13.5) with using a 60 second epoch, and a minimum wear-time of four days, with the inclusion of at least one weekend/non/workday as this has been shown to be a valid and reliable estimate of weekly sedentary behavior and physical activity level (58). Accelerometer data were processed by removing non-wear periods, trimming the start and end of recording, and checking for missing values in accordance with established guidelines (59). ActiGraph accelerometer physical activity data were processed utilising existing validated criteria and procedures (60). The Cole-Kripke algorithm (61) in ActiLife to examine sleep data (Total sleep time (TST), sleep efficiency (SE), sleep timing, regularity). Sleep periods were manually cross-referenced against participant sleep diaries and by visual inspection. This was conducted in line with the SBSM accelerometry guidelines (59). Data was omitted at the beginning and end of the recording when the device was not on the patient’s wrist. Any non-sleep periods detected as sleep (i.e. when device was taken off the wrist and not worn for a period of time) were removed - this was validated against the activity monitor logs and sleep diary. “In bed (or lights out)” and “out bed (wake up, time stopped trying to sleep)” times were compared between the ActiGraph with the sleep diary and were adjusted where necessary i.e. if the participant in the sleep log indicated they were awake, but lay in bed for another hour - this was adjusted on the ActiGraph data to reflect “out bed” time as time woke up in sleep diary. Instructions and an activity log were given for both devices where participants noted each time the device was removed for bathing or water activities.

A sleep diary was provided to participants to be completed over seven consecutive days, to track sleep patterns, monitor regularity and inform an accurate assessment of actigraphy recordings (62). The sleep diary was based upon the Consensus sleep diary and assessed various aspects of sleep, including sleep and wake time, sleep duration, and napping (62). The questions ask about: (1) the time of getting into bed; (2) the time at which the individual attempted to fall asleep; (3) sleep onset latency; (4) number of awakenings; (5) duration of awakenings; (6) time of final awakening; (7) final rise time; (8) perceived sleep quality (rated via Likert scale); and (9) an additional space for open-ended comments. An example of the sleep diary is given in Supplementary table 2.

### Venous blood sample collection

Capillary whole blood was collected using a sterile lancet from the medial or lateral aspect of the third, fourth or fifth finger. Capillary blood was then analysed instantly (AccuChek Instant S Meter Kit) to determine fasting blood glucose levels. Venous blood samples (20-30 mL) were then collected from the antecubital vein using a 21-gauge butterfly needle into prepared vacutainers (PAXgene, serum, and EDTA plasma) by a qualified phlebotomist. Samples were collected at a single time-point in the morning in a fasted state and stored on ice until preparation. After collection, serum samples were allowed to clot for 20-30 min at room temperature. Once serum was clotted, plasma and serum samples were centrifuged at 3000 rpm for 10 min at 4°C. Separated serum and plasma was aliquoted, boxed and then frozen in a –80 °C freezer before analysis. Blood collected in PAXgene tubes was stored in a -20 °C freezer before analysis.

### Blood pressure

An automatic digital blood pressure (BP) (OMRON) machine was used to assess BP. Participants were required to sit quietly for a minimum of 5 minutes prior to the measurement.

### Inflammatory, neurotrophic and bone factors

<u>Markers of inflammation</u> (leptin, adiponectin, TNF-α, IL-1β, IL-6, interleukin-8 (IL-8), interleukin-10 (IL-10), interferon-γ (IFN-γ), monocyte chemoattractant protein-1 (MCP-1)), bone turnover (osteopontin (OPN), and osteoprotergerin (OPG)), and neurotrophic factors (brain-derived neurotrophic factor (BDNF), and vascular endothelial growth factor A (VEGF-A)), will be ascertained from serum/plasma samples collected at assessment session two. Samples will be analysed using multiplex microsphere-based immunoassays (Luminex^®^, ThermoFisher).

DNA will be extracted using automated column-based techniques (Qiagen®) from PAXgene® tube samples and were utilised for whole genome sequencing (WGS), genome-wide association studies (GWAS), single nucleotide polymorphism (SNP) analysis for growth-related factors, DNA methylation studies and RNA expression studies.

### Hormonal factors

Markers of reproductive function (oestradiol (E2) and follicle stimulating hormone (FSH)) will be ascertained from blood serum collected from participants in assessment session two. Samples will be analysed using commercially available enzyme-linked immunoassays (Invitrogen). All blood biomarker analysis (Figure 3) will be conducted in the Molecular Biology laboratory, at the host institution.

**Figure 3.**
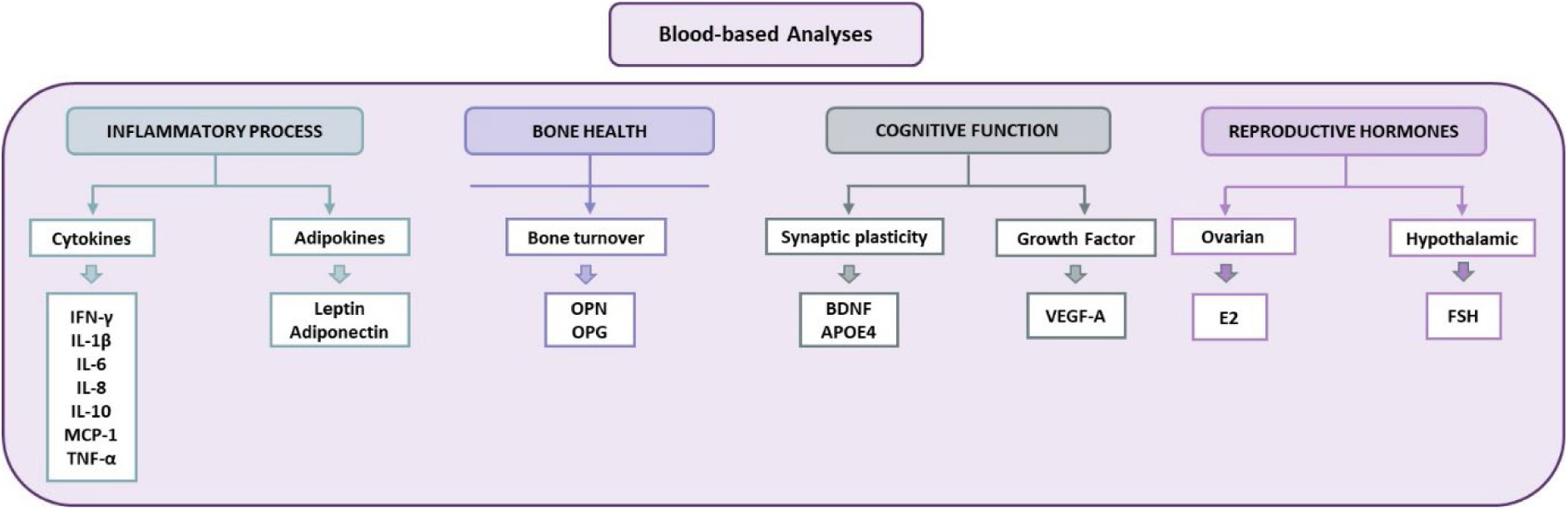
Blood-based biomarker assays to be completed under the LIFE protocol. Blood sample collection took place during the second data collection session and all blood biomarker analysis was conducted in the Molecular Biology laboratory, at the host institution. These included markers of inflammation (TNF-α, IL-1β, IL-6, IL-8, IL-10, IFN-γ, and MCP-1), markers of bone health (OPN and OPG), markers of cognitive function (BDNF and VEGF-A) and finally markers of reproductive function (E2 and FSH). TNF-α: Tumor necrosis factor-α, IL-1β: Interleukin-1β, IL-6: Interleukin-6, IL-8: Interleukin-8, IL-10: Interleukin-10, IFN-γ: Inteferon-γ, MCP-1: Monocyte chemoattractant protein-1, OPN: Osteopontin, OPG: Osteoprotegerin, BDNF: Brain derived neurotrophic factor, VEGF-A: Vascular endothelial factor-A, E2: Oestradiol and FSH: Follicle stimulating hormone.

### Physical performance

Dominant and non-dominant hand grip strength (HGS) was assessed using a spring-loaded grip dynamometer (TTM, Tokyo, Japan) to estimate physical performance and muscular strength (63). Participants performed a maximal contraction with alternating hands maintaining a 90-degree elbow flexion and limiting accessory movements. A brief rest period (approximately 10 seconds) was provided between subsequent attempts (63). The average of three measurements was taken as one assessment.

The 30-sec sit to stand protocol was used to assess functional leg strength. Participants were asked to sit in a hard-backed chair, with a seat height of 43cm from the floor, with their arms folded across their chest. Participants were instructed to rise as fast as possible to a full standing position then return to a full sitting position without using their arms (64). The assessor used a standardized script and provided balance support if necessary.

Balance was measured over three postures, tandem, semi-tandem and single leg balance. Participants were provided with an explanation and demonstration of each position and asked to balance each progressive position for 30 seconds with their eyes open. Participants were provided with two attempts at each position and the best time at each stage was recorded to the nearest 0.1 second (65). Participants were provided balance support until stable, after which timing commenced.

The Timed Up and Go test (TUG) is a validated measure used to assess walking ability and mobility in older adults (66). Starting in a seated position, participants were instructed to rise upon the researcher’s command “Go”, walk three meters forward (marked), turn around, walk back to the chair and sit down. Using a stopwatch, the research staff measured the time taken from their instruction to stand until the participant sat down again.

The 6-minute walk test is a practical measure frequently used in clinical settings as a sub-maximal test to assess functional exercise capacity (67). A 30 m track was marked in an under covered area outdoors. Using a stopwatch, the research staff instructed the participant to walk as far as they could in 6-minutes, with the number of laps completed and time recorded by the researcher. The total distance travelled during this time was then calculated.

### Neuropsychological testing

The Addenbrooke’s cognitive examination (ACE-III) is a brief, paper-based cognitive test that assesses multiple domains of cognitive function including attention, memory, fluency, language, and visuospatial abilities (42). A trained member of the research team asked the participant a series of set questions and instructions in a quiet room. The test took approximately 15 minutes to complete. The neuropsychological testing and corresponding cognitive domains are detailed in Figure 4.

**Figure 4.**
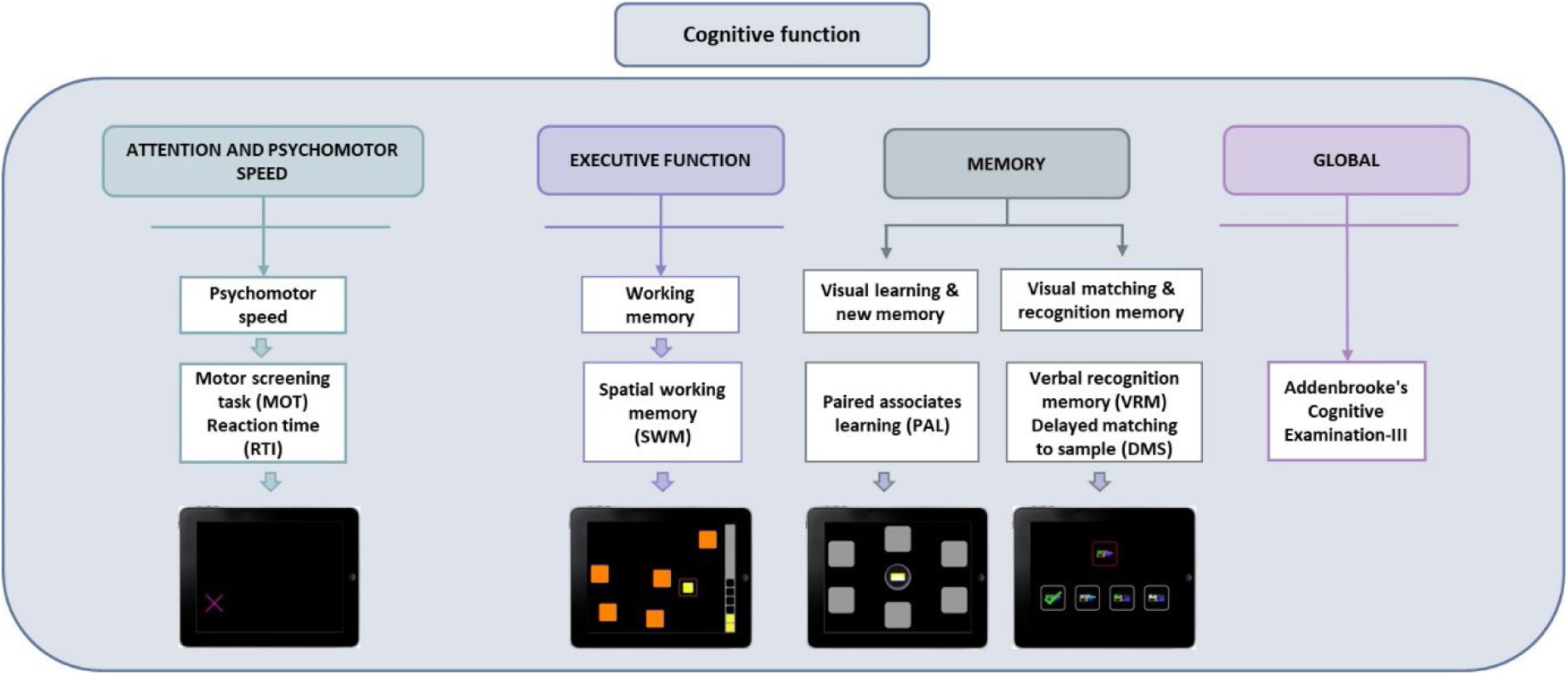
Neuropsychological testing and corresponding cognitive domains. Cognitive function assessment took place in the third data collection session. Global cognitive function was assessed with the Addenbrooke’s Cognitive Examination-III (ACE-III). Cambridge Automated Neuropsychological Test Assessment Battery (CANTAB) was used to assess the cognitive domains of executive function, attention and psychomotor speed and memory.

The Cambridge Automated Neuropsychological Test Battery (CANTAB) (68) is a computerised battery of cognitive assessments used to evaluate a range of cognitive functions including attention, psychomotor speed, executive function, and memory. CANTAB was self-administered using an iPad touch screen interface and took approximately 50 minutes to complete. Participants were asked to follow the audio instructions played through the application to perform the Motor Screening Task (MOT), which was used to introduce the CANTAB touchscreen to participants. The MOT provides a general assay that tests whether sensorimotor or other difficulties limit the collection of valid data from each participant. After touchscreen adaptation, the following CANTAB tests were completed, Paired Associates Learning (PAL), Spatial Working Memory (SWM), Delayed Matching to Sample (DMS), verbal recognition memory (VRM), and reaction time (RTI). CANTAB outcomes were automatically recorded in the application for later retrieval by study staff. All access to the application is controlled by the Licensee and access is granted via a password-controlled interface.

### Questionnaire measures

Questionnaires were administered during the testing session or emailed to the participants for self-completion. The RAW, MENQOL (37), Active Australia (39), PSQI (40), Audit-C (38), and family disease history questionnaire were combined and completed in assessment session one online. The remaining questionnaires (Depression and Anxiety Stress Scale (DASS)-21, activities balance confidence (ABC), Berlin Questionnaire, RUSATED, Insomnia Severity Index (ISI), Pre-sleep Arousal Scale (PSAS) and chronotype preference were administered together during assessment session three.

The DASS is a 21-item self-report instrument designed to measure the presence and severity of three related negative emotional states-depression, anxiety and stress - over the past 7 days (69). Participants were asked to respond to each item on a scale ranging from 0-3 (i.e., 0 being ‘did not apply to me at all’ and 3 being ‘applied to me very much, or most of the time). The DASS is clinically validated in measuring emotional states of depression, anxiety, and stress in clinical populations (44, 70).

To assess the falls risk of each participant, the ABC scale was completed at all testing time points (49). The ABC scale is a 16-item scale that requires participants to answer questions relating to their balance confidence when performing activities at home or in areas external to home. Each item is rated from 0% (no confidence) to 100% (complete confidence). The answers were summed and then divided by 16 to obtain a total ABC score. The ABC scale has been shown to have strong test-retest reliability, internal consistency and validity (49).

### Self-reported physical activity, sitting time and sleep

The Active Australia Questionnaire was used to measure the number of sessions and time spent per week engaged in walking, vigorous gardening/yard work, and vigorous physical activity (e.g., recreational sports, weight training). Participation in physical activity, measured as number of sessions and time per week for each question were calculated (39). Sitting time and activity was assessed using the Sedentary, Transport and Activity Questionnaire (71). Participants reported their sitting time over the week prior, categorized by weekdays and weekends, and across five contexts: work, transport, TV, viewing, leisure time computer use and ‘other’ sitting (71).

The 24-item PSQI assesses subjective sleep quality and disturbances in the preceding month. Five items, intended for a bed partner or roommate provided descriptive value only. The other 19 items are divided into seven “component” scores (subjective sleep quality, sleep latency, duration, efficiency and disturbance, use of sleep medication and daytime dysfunction), each of which has a range of 0-3 points.

In all cases, a score of “0” indicates no difficulty, while a score of “3” indicates severe difficulty. The seven-component scores are then added to yield a “global” score, with a range of 0-21 points, “0” indicating no difficulty and “21” indicating severe difficulties (40).

The ISI is a seven-item measure of the nature, severity, and impact of current (i.e., past 2 weeks) insomnia symptoms, with participants responding on a Likert-type scale (i.e., 0 being ‘not at all’ to 4 being ‘very much’). The total score ranges from 0 to 28, with a higher score indicating more severe insomnia (72). The questions relate to the subjective qualities of the respondent’s sleep, including satisfaction with sleep patterns, the degree to which insomnia interferes with daily functioning, and whether symptoms are noticeable to others (72).

RUSATED is a self-administered questionnaire evaluating six key dimensions of sleep health shown to be associated with various health outcomes: 1) regularity (‘do you go to bed and get out at the same times [within one hour] every day?’, 2) satisfaction (‘are you satisfied with your sleep?’), 3) alertness (‘do you stay awake all day without dozing?’), 4) timing (‘are you asleep [or in bed] between 2.00am and 4.00am?’), 5) efficiency (‘do you spend less than 30 minutes awake at night?’), and 6) duration (‘do you sleep between 6 and 8 hours per day’). Each item is rated on the frequency of meeting the criteria for each dimension from 0 to 2, with 0 for “never” or “rarely,” 1 for “sometimes,” and 2 for “usually” or “always.” (73)

The Berlin Questionnaire (BQ) assesses the degree of risk for obstructive sleep apnoea, incorporating questions about snoring (category 1), daytime somnolence (category 2), and hypertension and BMI (category 3). The overall BQ score is determined, from the responses to the three categories: scores from the first and second categories were positive if the responses indicated frequent symptoms (>3–4 times/week), whereas the score from the third category was positive if there was a history of hypertension or a BMI >30 kg/m^2^. Patients were scored as being at high-risk for obstructive sleep apnoea if they had a positive score on two or more categories, while those who did not were scored as being at low-risk (46).

The Pre-sleep Arousal Scale (PSAS), is a 16-item self-report questionnaire designed to assess arousal levels before falling asleep (ref). It consists of two subscales, these being cognitive and somatic, both demonstrating adequate internal consistency (α=0.82 and 0.79, respectively) (48). The PSAS is rated on a 5-point Likert scale ranging from 1 (‘not at all’) to 5 (‘extremely’) with total scores for each subscale ranging from 8 to 40. Higher scores indicate greater pre-sleep arousal (48).

The self-morningness/eveningness (Self-ME) is a single item scale designed to assess chronotype preference, “How would you describe yourself” (i.e. 1) Definitely a morning person, 2) More a morning than an evening person, 3) Both morning and evening person, 4) More an evening than a morning person, 5) Definitely an evening person, 6) Don’t know), which has previously validated (74).

### Dietary intake

Caffeine use was determined through a single question asking “Thinking about caffeinated beverages such as energy drinks, soft drinks, coffee and tea, how many cups or cans of caffeinated beverages do you typically drink each day? Responses ranged from 1-10 or more or refuse to answer. Habitual dietary intake was assessed using the Australian Eating Survey (AES), a validated 120-item semi-quantitative food frequency questionnaire (75) with 15 supplementary questions regarding age, use of vitamin supplements, food behaviors and sedentary behaviors. The AES is a self-administered online tool assessing dietary intake over the preceding 6 months. The tool was administered using an iPad during assessment session three. An individual response for each food, or food type, was required, with frequency options for food ranging from ‘never’ to ‘4 or more times per day’, dependent on the food, and up to ‘7 or more glasses per day’ for some beverages. Nutrient intakes are computed from the Australian food composition database AusNut 2011-2013 primarily and AusFoods (Brands) Revision 5 (Australian Government Publishing Service, Canberra).

### Socio-demographic characteristics and clinical information

Sociodemographic characteristics were obtained as part of the RAW questionnaire in assessment Session 1 which included factors such as level of education, employment status, ethnicity, alcohol consumption (AUDIT-C) and smoking status. Full details are given in Supplementary table 3.

## DATA MANAGEMENT PLAN

All data collection procedures were conducted according to the study Standard Operating Procedure (SOP) Manual. All research staff involved in a data collection procedure were appropriately trained and assessed in competence by the Principal Investigator. Inter- and intra-tester reliability was assessed for all individuals involved in the data collection procedure. The Principal Investigator performed quality control activities to verify study data and ensure data was complete and accurate. In addition, study-related procedures and processes, including adherence to SOP’s was monitored to ensure alignment with the approved protocol, good clinical practice (GCP) and Institutional guidelines. The study case report forms (CRF) were the primary data collection instruments for this study, therefore the accuracy, completeness, legibility and timeliness of the data reported was essential. All data requested on the CRF had to be recorded, missing data were explained. Error entries were corrected by drawing a single straight line through the incorrect entry so as not to obliterate the original entry and the correct data was entered above it. Changes were initialled and dated.

Information about the participants in the trial was obtained and stored in paper and electronic file format throughout and after the study. In accordance with Good Clinical Practice (GCP) Guidelines the trial files and documents were kept in a secured filing area The storage area is locked and accessible to staff who have security access. All electronic databases are password protected. Reported study data are verifiable from the source documents. Source data includes all information, original records of clinical findings, observations, or other activities in clinical trials necessary for the reconstruction and evaluation of the trial. Source data are contained in the original source documents.

## SAFETY CONSIDERATIONS

There are no significant harms or risk associated with participation in the LIFE study. The procedures used were designed to have minimal risk to participants. Participants may have experienced some minor and transient issues with the assessments, which are described below:

DXA is a routine measurement for bone density and body composition and the associated dose with DXA (GE Lunar iDXA) was 3 µSv for the whole body. In comparison, an individual receives between approximately 4-5.5 µSv for daily natural background exposure, 80 µSv for a return trans-Pacific flight and 100 µSv for a chest x-ray. Therefore, although ionising radiation was used in the scan the corresponding risk from participating in this study is low. In addition, assessment and approval for the use of DXA ionising radiation from the School of Health and Behavioral Sciences Local Radiation Safety Officer and The University of the Sunshine Coast Radiation Safety Officer was obtained prior to commencement of this study.

All venous blood sampling procedures were carried out by a qualified phlebotomist to minimise risk of bruising, and all occupational health and safety procedures were followed to minimise the risk of infection associated with venous blood sampling. All blood sampling procedures were carried out in dedicated laboratory spaces.

The risk of misuse of genetic information derived from this study is very low. Breach of confidentiality could occur during data collection or the biological sample collection procedures. Data and biological samples will be stored in accordance with the Australian Privacy Guidelines (April 2014) and the NHMRC National Statement on Ethical Conduct in Human Research (2007).

Completing questionnaires may be stressful for some participants depending on their cognitive status, mood, willingness etc. Care was taken when administering questionnaires and if any stress or discomfort was observed the participant was given the option of returning to the test/questionnaire at a later time.

Clinical laboratory abnormalities were to be documented as an adverse event if any one of the following conditions were met: a) the laboratory abnormality is not otherwise refuted by a repeat test to confirm the abnormality. b) the abnormality suggests a disease and/or organ toxicity; and the abnormality is of a degree that requires active management, e.g. more frequent follow-up assessment, further diagnostic investigation by a medical professional etc.

Questionnaire values were to be documented as adverse events if any of the following conditions were met a) Extremely high and low scores were monitored and discussed with the participant for appropriate referral to their treating medical practitioner. b) If any anxiety or stress was caused from completing any of the study surveys this was to be documented as an adverse event and the participant recommended to visit their general practitioner.

Information on all adverse events were to be recorded immediately in the source document and also in the appropriate adverse event module of the case report form. All clearly related signs, symptoms, and abnormal procedural results were to be recorded in the source document. The clinical course of each event was to be followed until resolution, stabilisation, or until it has been determined that the study intervention or participation is not the cause.

All serious adverse events were to be reported within 24 hours of occurrence, in accordance with the study protocol and GCP Guidelines. A serious adverse event is an inability to carry out usual activities and were to be reported to the HREC through the appropriate pathways. The investigator kept a copy of the procedure on file at the study site.

## STATISTICAL ANALYSIS PLAN

Data will be analysed using SPSS (version 22.0, SPSS, Inc., Chicago IL USA) and Prism^®^ (Version 7.0, GraphPad, Inc., San Diego CA USA) statistical software packages. Data will be analysed per protocol, and participants with missing data points will be excluded from formal analysis. General linear models including ANOVA, linear regression and generalised linear models (where appropriate) will be used to address study aims. All assumptions will be tested, and statistical significance will be set at p<0.05.

## DATA AVAILABILITY

Data supporting the findings of this study will be made available upon request to the corresponding author. The authors confirm that the supporting data and any subsequent findings from this study will be available within published articles and supplementary materials

## PILOT RESULTS

Recruitment for the LIFE study commenced in October 2022 and data collection was completed in May 2024. Data analysis is ongoing and is expected to be completed by end of 2026. Participant characteristics are summarized in Table 2. Participants (n=165) are distributed across premenopausal (n=43), perimenopausal (n=55), and postmenopausal (n=67) stages. The original target sample size of n=175 calculated from the power level of 0.95 was not reached. However, a sample size calculation with power level reduced to 0.90, alpha level of 0.05, determined n=144 (48 per group) was sufficient. Although the number of participants exceeds 144 it is of note that the original sample size was not reached when discussing future analysis and results.

**Table 2.** Participant characteristics.

|  | **All (n=165)** | **Pre (n=43)** | **Peri (n=55)** | **Post (n=67)** |
| --- | --- | --- | --- | --- |
| **Age** mean (SD) | 51.30 ± 6.82 | 45.04 ± 3.29 | 48.12 ± 3.69 | 57.93 ± 4.36 |
| **Ethnicity** n (%) |  |  |  |  |
| Australian | 143 (87) | 34 (79) | 46 (84) | 63 (94) |
| Aboriginal/Torres Strait Islander | 3 (2) | 2 (5) | 1 (2) |  |
| New Zealander | 4 (2) | 2 (5) | 1 (2) | 1 (1) |
| North-West European | 16 (10) | 7 (16) | 7 (13) | 2 (3) |
| Southern and Eastern European | 1(1) |  |  | 1 (1) |
| North-East Asian | 2 (1) | 1 (2) | 1 (2) |  |
| Peoples of the America's | 3(2) | 2 (5) |  | 1 (1) |
| Other | 6 (4) | 1 (2) | 3 (5) | 2 (3) |
| **Marital status** n (%) |  |  |  |  |
| Married or partnered | 132 (80) | 36 (84) | 44 (80) | 52 (78) |
| Single (never married) | 15 (9) | 1 (2) | 7 (13) | 7 (10) |
| Separated or divorced | 16 (10) | 6 (14) | 4 (7) | 6 (9) |
| Widowed | 2 (1) |  |  | 2 (4) |
| **Household** n (%) |  |  |  |  |
| My partner/spouse | 125 (76) | 34 (79) | 39 (71) | 52 (78) |
| My children | 95 (58) | 40 (93) | 41 (76) | 14 (21) |
| Parent(s) | 7 (4) | 1 (2) | 1 (2) | 5 (7) |
| Other | 6 (4) |  | 2 (4) | 4 (6) |
| Live alone | 12 (7) |  | 3 (5) | 9 (13) |
| **Education level** n (%) |  |  |  |  |
| Bachelor degree or higher | 94 (57) | 28 (65) | 39 (71) | 27 (40) |
| Certificate/diploma | 44 (27) | 11 (26) | 10 (18) | 23 (34) |
| Left school after age 16 | 10 (6) | 2 (5) | 2 (4) | 6 (9) |
| Left school at 16 years or less | 8 (5) |  | 1 (2) | 7 (10) |
| Other | 9 (6) | 2 (5) | 3 (5) | 4 (6) |
| **Employment status** n (%) |  |  |  |  |
| Working full-time | 56 (34) | 17 (40) | 22 (40) | 17 (25) |
| Working part-time | 58 (35) | 16 (37) | 22 (40) | 20 (30) |
| Working more than one job | 17 (10) | 3 (7) | 8 (15) | 6 (9) |
| Student | 21 (13) | 11 (26) | 10 (18) | 4 (6) |
| Homemaker | 19 (12) | 10 (23) | 6 (11) | 3 (4) |
| Unemployed | 1 (1) |  |  | 1 (1) |
| Retired | 9 (6) |  |  | 9 (13) |
| Volunteer | 6 (4) | 2 (5) | 1 (2) | 3 (4) |
| Other | 22 (13) | 3 (7) | 4 (7) | 15 (22) |
| **Work schedule** n (%) |  |  |  |  |
| Regular day shifts | 102 (62) | 30 (70) | 39 (71) | 33 (49) |
| Regular evening shifts | 1 (1) |  | 1 (2) |  |
| Rotating night shifts | 2 (1) | 1 (2) |  | 1 (2) |
| Other | 20 (12) | 4 (9) | 6 (10) | 8 (12) |
| **Smoking status** |  |  |  |  |
| Current smoker | 2 (1) |  | 1 (2) | 1 (2) |
| No, but used to smoke | 53 (32) | 14 (33) | 16 (29) | 23 (34) |
| No, never smoked habitually | 110 (67) | 29 (67) | 38 (69) | 43 (64) |
| **Alcohol intake** |  |  |  |  |
| Never | 14 (9) | 1 (2) | 9 (16) | 4 (6) |
| Monthly or less | 33 (20) | 13 (30) | 10 (18) | 10 (15) |
| 2-4 times per month | 33 (20) | 10 (23) | 11 (20) | 12 (18) |
| 2-3 times per week | 51 (31) | 12 (28) | 20 (36) | 19 (28) |
| 4+ times per week | 34 (21) | 7 (16) | 5 (9) | 22 (33) |

The majority of participants identify as Australian (87%), with smaller proportions identifying as European (10%), New Zealander (2%), Peoples of the Americas (2%), Aboriginal and / or Torres Strait peoples (2%) and other ethnic groups (1% each for Asian and Southern and Eastern European). Most participants are married or partnered (80%) and live with their spouse (76%), with similar proportions across groups. The majority of participants have either a Bachelor degree or higher (57%), although there is some difference in education level between pre- (65%), peri- (71%) and post- (40%) menopause groups. Employment data revealed that most participants are either working full or part time (34% and 35% respectively), with similar distributions across groups aside from the number of participants who identified as “Homemaker” (pre - 23%, peri – 11% and post – 4%). This is most likely due to the average age of participants across the groups. The majority of participants have never habitually smoked (67%) and drink alcohol between 2-4+ times a week (52%). There were no adverse events reported in this study.

### Implications for future research

The primary objective of the LIFE study is to develop an understanding of how hormonal and lifestyle behavior differences in pre, peri and postmenopausal women influence chronic systemic inflammation and visceral adiposity to drive future research in the prevention of chronic inflammatory conditions and direct future lifestyle interventions for older women at risk of obesity-related chronic disease.

The secondary objectives are to firstly, identify and compare associations of chronic systemic inflammation and visceral adiposity, on cognitive function and wellbeing (depression, anxiety and stress measures) between pre, peri and postmenopausal women. Secondly, to identify the relationship between lifestyle behaviors and sleep measures and menopausal symptoms. Investigating the underlying mechanisms linking unfavourable differences in body composition, chronic inflammation, and cognitive function will assist in the development of future strategies to mitigate chronic disease risk and prevent cognitive decline in older women.

Collectively these objectives providing an understanding and characterization of lifestyle behaviors and links to inflammatory markers, cognition and sleep health in pre, peri and postmenopausal women will inform targeted strategies to improve wellbeing, heart, brain, and metabolic health long-term.

## Acknowledgments

The authors would like to thank all the participants who volunteered their time to partake in this study. The authors acknowledge the contributions of additional LIFE study team members and students; Arshia Kaur, Cintia Carvalho, Corey Linton, Jasmin Elliott, Amyliah Harrison, Abigail Sidey, and Isabella Buckland for their contribution to recruitment and data collection.

