## Supplemental material for "The LIFE Study: a cross-sectional study protocol for LIfestyle risk Factors for chronic disease across the stagEs of reproductive ageing"

**SUPPLEMENTARY MATERIAL**

**Supplementary table 1.** Eligibility criteria

| **Inclusion** | - Cisgendered women or non-binary   *Cisgendered refers to a person whose sex assigned at birth and gender identity are the same. For the purpose of this research study, non-binary includes those whose sex at birth was female and identity is non-binary.*   - Aged between 40-65 years - Body mass index (BMI) 18.5– 39.9kg/m^2^ - Able and willing to provide informed consent - Able to communicate in English - No history of stroke or brain trauma - No presently diagnosed mental illness or significant cognitive impairment - Stable body weight (± 5%) for > 3 months prior to beginning the study (self-report) |
| --- | --- |
| **Exclusion** | - Pregnant or breastfeeding - Exogenous hormone use including but not limited to hormone replacement therapy, oral contraceptive use, and intrauterine device. - Treatment/surgically induced menopause - Diagnosed hormonal or ovarian disorder (polycystic ovarian syndrome, hypothalamic amenorrhea) - Conditions that may influence outcomes of the research study, such as uncontrolled inflammatory conditions. - Major illness/physical problems (acute or chronic) that may limit participation (e.g., liver, renal, or haematological disease) - Evidence of untreated hypertension (blood pressure >160/90mmHg) and/or hypertriglyceridemia (self-report) - Significant cardiovascular disease (unstable angina, cardiac failure) or recent myocardial infarction, coronary artery bypass graft, ischaemic or haemorrhagic stroke in the previous 3 months. - Cancer diagnosis/treatment in the previous 12 months - A medially diagnosed and untreated severe sleeping disorder - Participants with known internal artefacts that may obscure DXA readings. |

**Supplementary table 2.** Recommendations from Student Equity and Diversity Advisor, University of the Sunshine Coast.

| Recommendation: | Action: |
| --- | --- |
| Use term ‘cis-gendered women’ to attract people whose sex at birth aligns with their gender identify.  Add definition. | Study material to include ‘cis-gendered women/woman’ as part of inclusion criteria.  Definition ‘cisgendered is a person whose sex assigned at birth and gender identity are the same’ added to all recruitment and study materials where the term cisgendered is used. |
| Add additional wording to include those who identify as non-binary. | Study material to include ‘non-binary’, with definition ‘for the purpose of this study, non-binary refers to women whose sex at birth was female and gender identity is non-binary'. |

**Supplementary Material 3.** Sleep diary
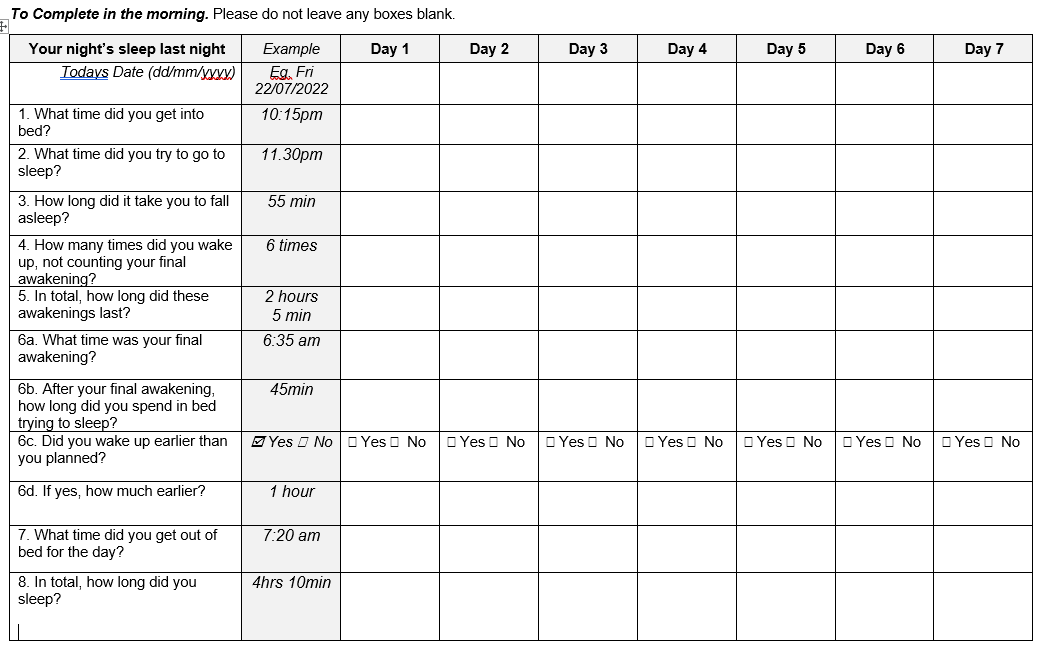

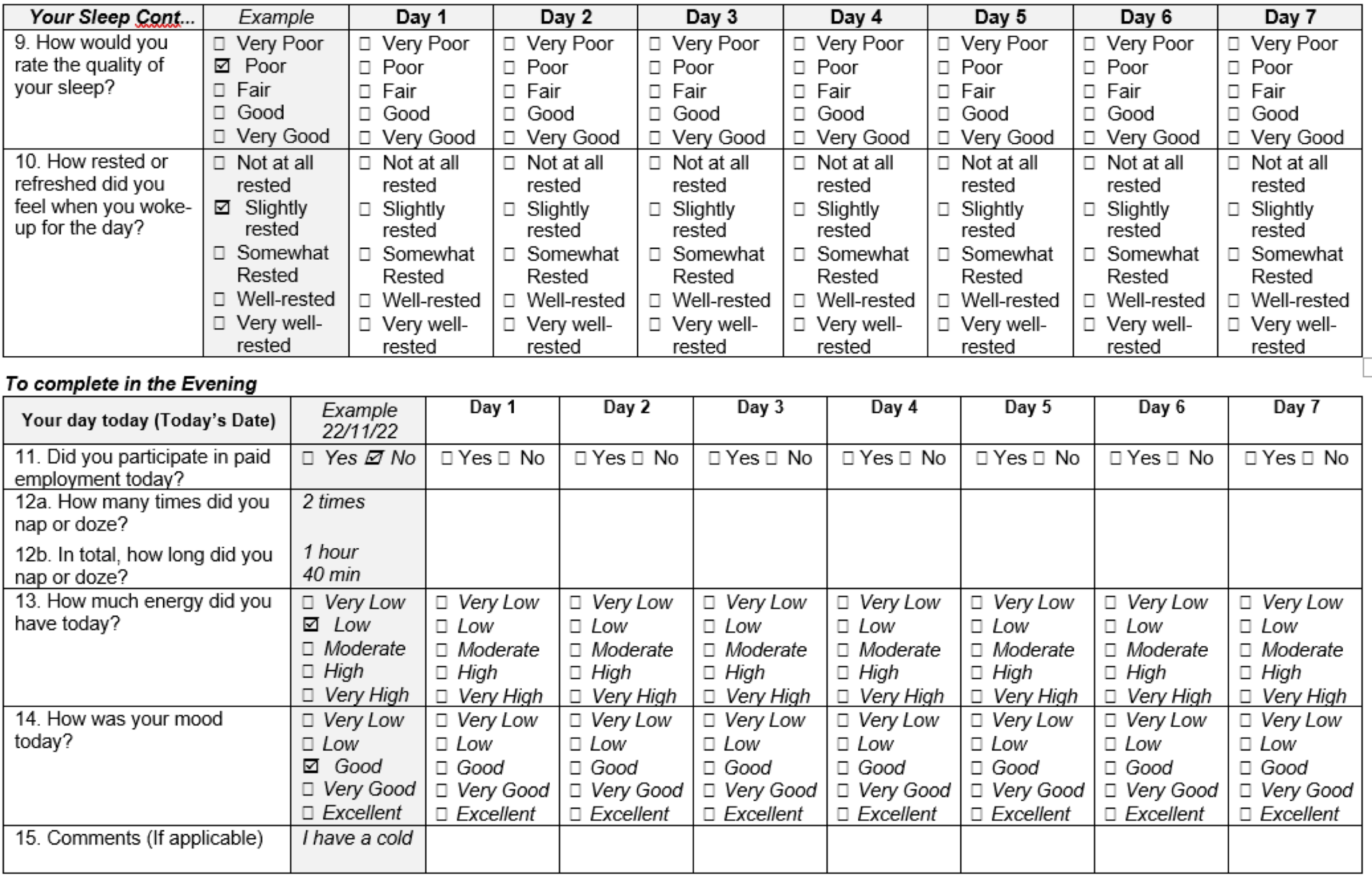
